# Challenges in conducting sexual health and violence research in older adults beyond GDPR: a Belgian case study

**DOI:** 10.1101/2020.09.18.20197350

**Authors:** Anne Nobels, Adina Cismaru Inescu, Laurent Nisen, Bastien Hahaut, Gilbert M.D. Lemmens, Christophe Vandeviver, Ines Keygnaert

**Affiliations:** International Centre for Reproductive Health, Department of Public Health and Primary Care, Ghent University, Ghent, Belgium; CARE-ESPRIst, Études et évaluations, University of Liège, Liège, Belgium; Department of Psychiatry, Ghent University Hospital, Ghent, Belgium; Department of Criminology, Criminal Law and Social Law, Ghent University, Ghent, Belgium; Research Foundation—Flanders (FWO), Brussels, Belgium

**Keywords:** Privacy, sampling, methodology, ageing, sexuality, elder abuse and neglect

## Abstract

**Background:** Because of a growing older population, the sexual health (SH) of older adults, including sexual violence (SV), is becoming an increasingly important public health concern. Yet, reliable SV prevalence rates and risk factors are lacking, due to methodological shortcomings in current studies. SV research involves challenges regarding safety and disclosure, especially in older adults. In this paper we reflect on the methods used in a SH&V study in older adults balancing between GDPR imposed privacy rules and ethical and safety guidelines.

**Methods:** To ensure the acceptability of the questionnaire, it was tested in a two-phase pilot study. To maximize SV disclosure, the questionnaire built up gradually towards the more sensitive SV modules. Interviewers were trained to approach participants in a non-judgemental manner. Due to GDPR, our data collection method was changed from a random sampling via the National Register to a cluster random probability sampling with a random route finding approach.

**Results:** Older adults were willing to discuss SH&V during a face-to-face interview with trained interviewers. Following strict safety guidelines, no major incidents were reported. The cluster random probability sampling with random route finding approach provided an adequate sampling frame, but was inefficient and time-consuming.

**Conclusion:** Doing research on SH&V in older adults is feasible, but requires a substantial investment of time and the challenges involved may incur greater costs. Research institutions, donors, and policy makers should convene to investigate how problems related to GDPR can be solved, especially regarding research on sensitive topics and hard to reach populations.

## INTRODUCTION

Since the older population in Europe and Belgium is expanding [1, 2], the sexual health (SH) of older adults, including sexual violence (SV), is becoming an increasing public health concern. According to recent meta-analysis, 0.9% of older adults and 2.2% of older women worldwide were sexually victimised in the past year [3, 4]. Life time SV prevalence was estimated at 6% [5]. However, these numbers are likely to be underestimated because of methodological shortcomings. Therefore, reliable SV prevalence numbers and associated risk factors in older adults are currently unavailable [6].

When doing SV research, protecting the participants and interviewers from potential violence by the assailant is of the utmost importance. When the topic of the study becomes known – either within the household or in the wider community-the assailant may find out the nature of the study, leading to possible safely issues for the participant or interviewer. Therefore, the World Health Organization (WHO) developed ethical and safety recommendations for SV research [7]. Also, as victims experience many barriers for SV disclosure [8], study designs should facilitate disclosure. This could be extra challenging in older adults, as society considers them asexual [9-11]. Older adults may internalise this stereotypical societal image of ‘the asexual older adult’, impacting SV disclosure [12]. In addition, discussing SH and SV with older adults is considered inappropriate [13, 14].

Further, since the 25^th^ of May 2018, the General Data Protection Regulation (GDPR) (EU) 2016/679 [15] imposed strict rules regarding collecting, storing and accessing personal data. In this study the implementation of the GDPR has led to an adaptation of the data collection procedure. In the original study design the Belgian National Register (NR) would serve as the sampling frame. This sampling frame closely overlaps with the target population and contains information on all Belgian residents [16]. Yet, since the implementation of GDPR, the NR only shares personal details of possible participants of scientific research via an active opt-in procedure. This implied drastic changes in the study protocol of this population-based study on SV in older adults.

In this paper we describe the methodology of the first SV prevalence study in older adults in Belgium. We reflect on the challenges in conducting research on a sensitive topic in a hard to reach population keeping the balance between the privacy rules imposed by the GDPR and the ethical and safety guidelines for violence research. Based on our experiences, we formulate recommendations for future research and policies.

## METHODS

### Used SV definition

In this study we adopted the WHO definition of SV which includes sexual harassment, sexual abuse with physical contact without penetration and (attempted) rape [17, 18]. Based on recent insights in the field of SV in older adults, this definition was expanded with sexual neglect [6, 19].

### The questionnaire

The questionnaire development comprised a multistep process of discussion and consultation. The multidisciplinary research team developed a first draft of the questionnaire following an extensive review of literature and pre-existing study instruments [20-24]. This draft was reviewed by the expert steering committee consisting of national and international researchers, policy makers and practitioners in the field of SV or elder abuse and neglect. The questionnaire aimed to maximize SV disclosure. To that extent, the questionnaire started with questions on less sensitive topics and built up towards the modules on SV victimisation and perpetration. All questions on SV and its consequences were phrased in a supportive and non-judgemental manner [7]. To assess SV experiences, behaviour specific questions based on the Sexual Experience Survey (SES) [23] and the Sexual Aggression and Victimization Scale (SAV-S) [21] were used, which were adapted to the Belgian social and legal context.

In order to test the acceptability and feasibility of the questionnaire in older adults, we performed a two-phase pilot study. In the first face validity phase we measured the extent to which the questionnaire appeared relevant, important and interesting to our target population [25]. We discussed the questionnaire with 12 older adults and consulted ten experts working with older adults. Based on their recommendations, we added extra questions on family life and SH at the beginning of the questionnaire to ease respondents in and adapted the questions on rape myths and gender norms. This adapted draft and the applied data collection procedure were tested in the second phase of the pilot study. We performed 50 face-to-face interviews with community-dwelling older adults. Based on participants’ recommendations and interviewers’ experiences we deleted the module on gender norms and reduced the number of follow-up questions in the SV perpetration module. The final questionnaire consisted of 13 modules (see Figure 1).

**Figure 1:**
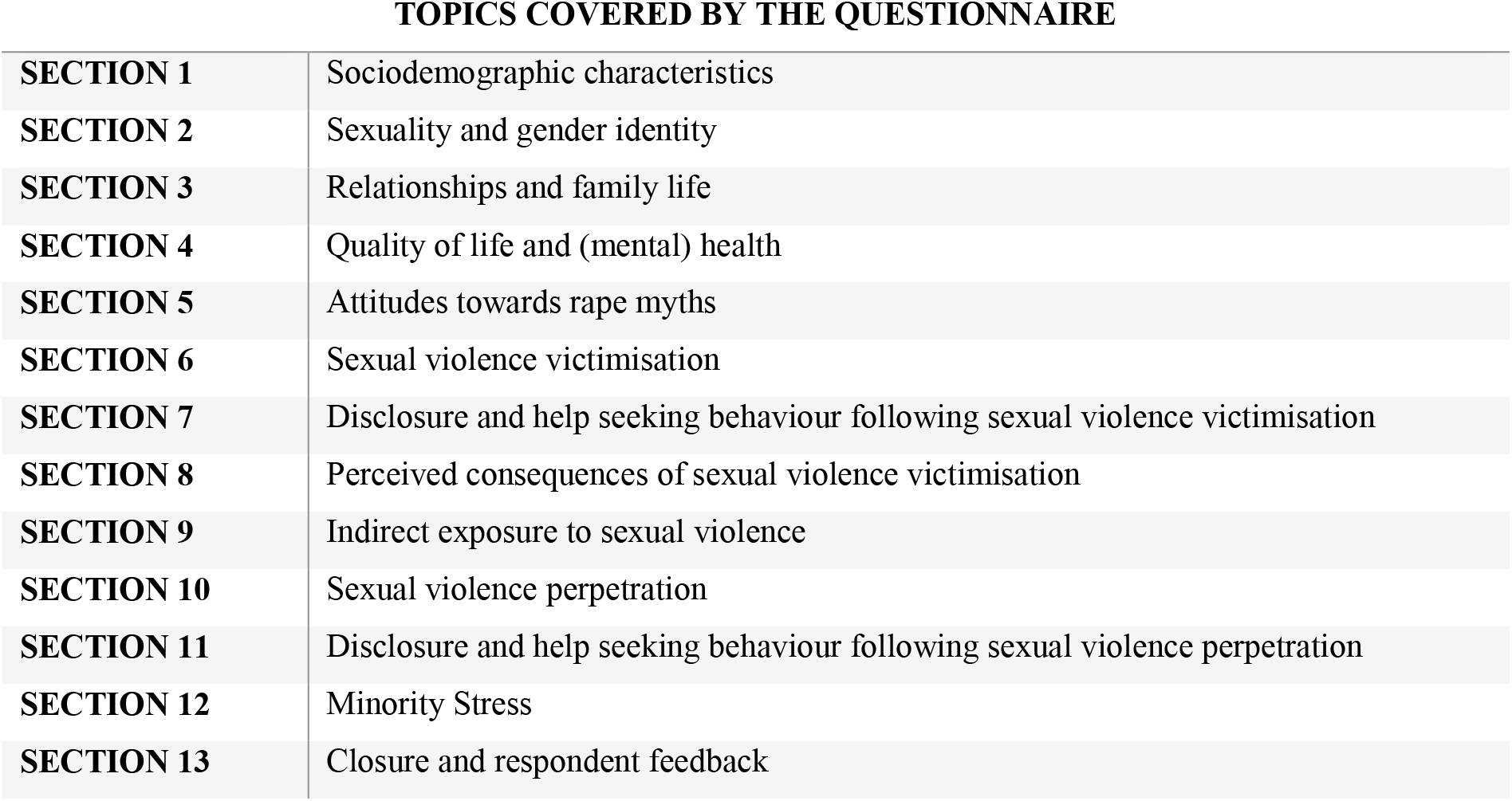
Flow of the questionnaire.

### Interviewers

Interviews were performed by the first author and 48 external interviewers (67% female, 33% male, mean age of 43 years). All interviewers were carefully selected and received specialised training [7]. During the selection process, specific attention was given to experience with older adults, social skills, experiences with and attitudes towards SV and coping mechanisms. After a first selection round, all interviewers participated in a multi-day training by the researchers and the coordinator of the study. Considering that rates of reported SV can be influenced by a suggestion of judgement or blame, the training confronted the interviewers with their own attitudes, fears and stereotypes towards SV and older adults and gave them the opportunity to come to terms with their own (direct or indirect) exposure towards SV [7]. The training comprised the following modules: SV (definition, prevalence and myths), discussing sensitive topics, communication skills, interview techniques, handling confidential information, and ethical and safety procedures. Interviewers had to familiarize themselves with the questionnaire during small group sessions, which helped them to feel at ease discussing SV.

Interviewers received close guidance from the research team. The researchers were always available to be contacted by phone or email to discuss difficulties experienced or questions during the fieldwork. Interviewers were instructed to call after the first day of interviewing and in the case of encountering a dangerous situation. In addition, all interviewers participated in at least two debriefing meetings. If necessary individual discussions with the researchers were arranged. Furthermore, we used a social media group in which interviewers indicated their progress, concerns and questions and a weekly newsletter, discussing the progress, pitfalls and recommendations for the fieldwork.

## Data collection

### Impact of GDPR on data collection procedure

Due to the GDPR, we changed our data collection procedure from a random sampling to a cluster random probability sampling with a random walk finding approach [26]. Originally, the intention was to acquire the contact details (name and address) of a representative sample of older adults living in Belgium through the National Register (NR). This sampling frame closely overlaps with the target population [16]. However, since the implementation of the GDPR, the NR can only share personal details of possible participants of scientific research via an active opt-in procedure. A representative sample of potential participants would be contacted by the NR, informing them of the study’s goals and asking them to provide their written consent and contact details to the research team. This procedure could lead to bias in participation, endangering the representativeness of the sample and the validity of the research results [27] as it encourages people with pronounced opinions on SV to participate [28] and excludes vulnerable older adults (e.g. older adults who cannot physically reach the mail box due to illness or disability). In addition, when participants need help to provide their written consent to the research team, there is a risk the assailant may find out the nature of the study, leading to safety issues for the participant [7]. Applying a cluster random probability sampling with random route finding approach, researchers don’t need contact details to contact older adults. Furthermore, the interviewer can assess the safety of the participant in real time when calling at their door.

### Applying the cluster random probability sampling

To guarantee precise and reliable estimates, 845 interviews needed to be performed (see Figure 2). Compared to the original procedure, the assumed design effect of the clustered sampling doubled the sample size of the study. A previous study shows that lifetime SV prevalence of Belgian older adults is 6% [5]. However, this rate is likely underestimated because of methodological shortcomings [6]. Since studies done in younger populations show a life time prevalence of SV between 10 and 30% [20-22], we assumed a 10% life time prevalence of SV in our sample (see Figure 2). Considering the spread of older adults is normally distributed on the municipality level in Belgium, 141 clusters of 6 interviews, geographically covering all regions, were randomly selected. Selection was based on the proportion of older adults of 70 years and older living in each municipality [29].

**Figure 2:**
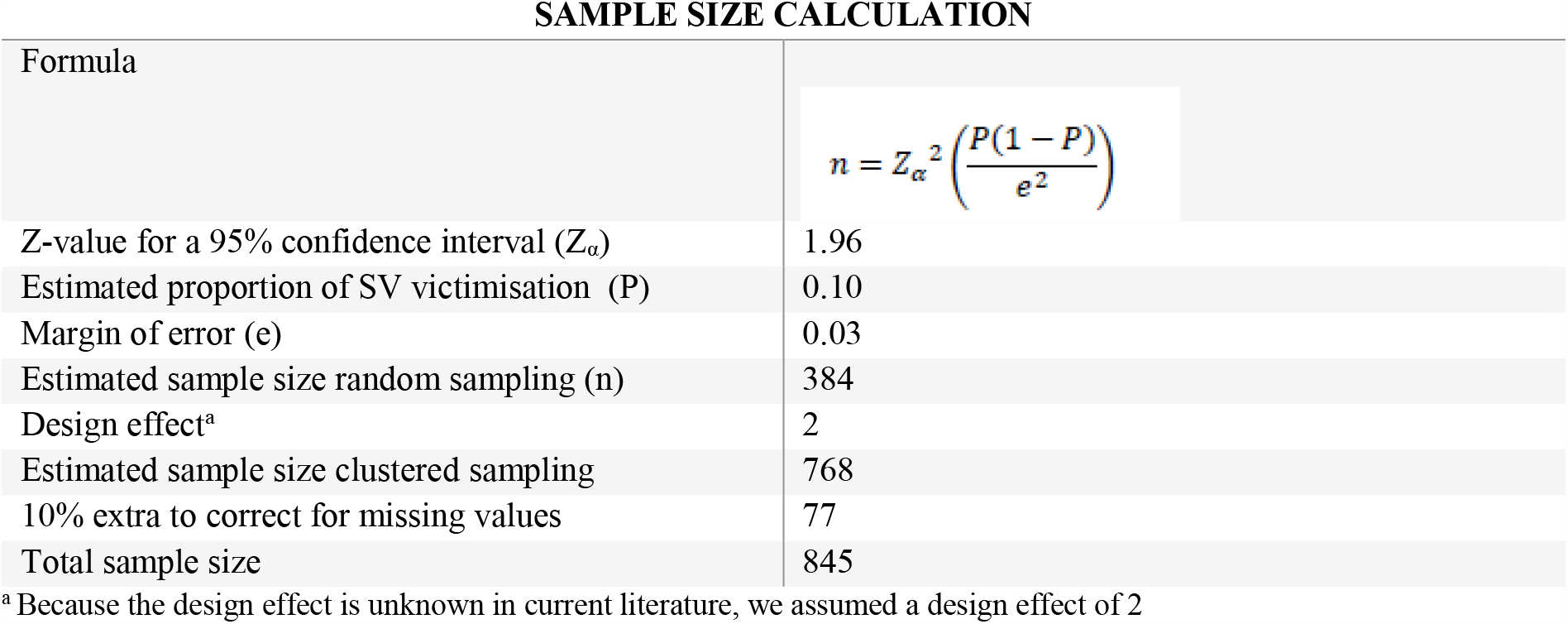
Sample size calculation.

Participants had to be 70 years or older, live in Belgium, and have sufficient cognitive ability to complete the interview. Both older adults living in the community and older adults living in nursing homes or assisted living facilities were included. Cognitive status was not formally assessed but was evaluated based on the ability to maintain attention during the interview and the consistency of the participant’s answer [30] by means of a control question comparing the participant’s birthyear and age. Eligible participants within each cluster were identified using a random walk finding approach [31]. Interviewers were provided with a randomly selected starting address and followed a strict set of rules that guided the selection of subsequent houses at pre-specified intervals (see Figure 3).

**Figure 3:**
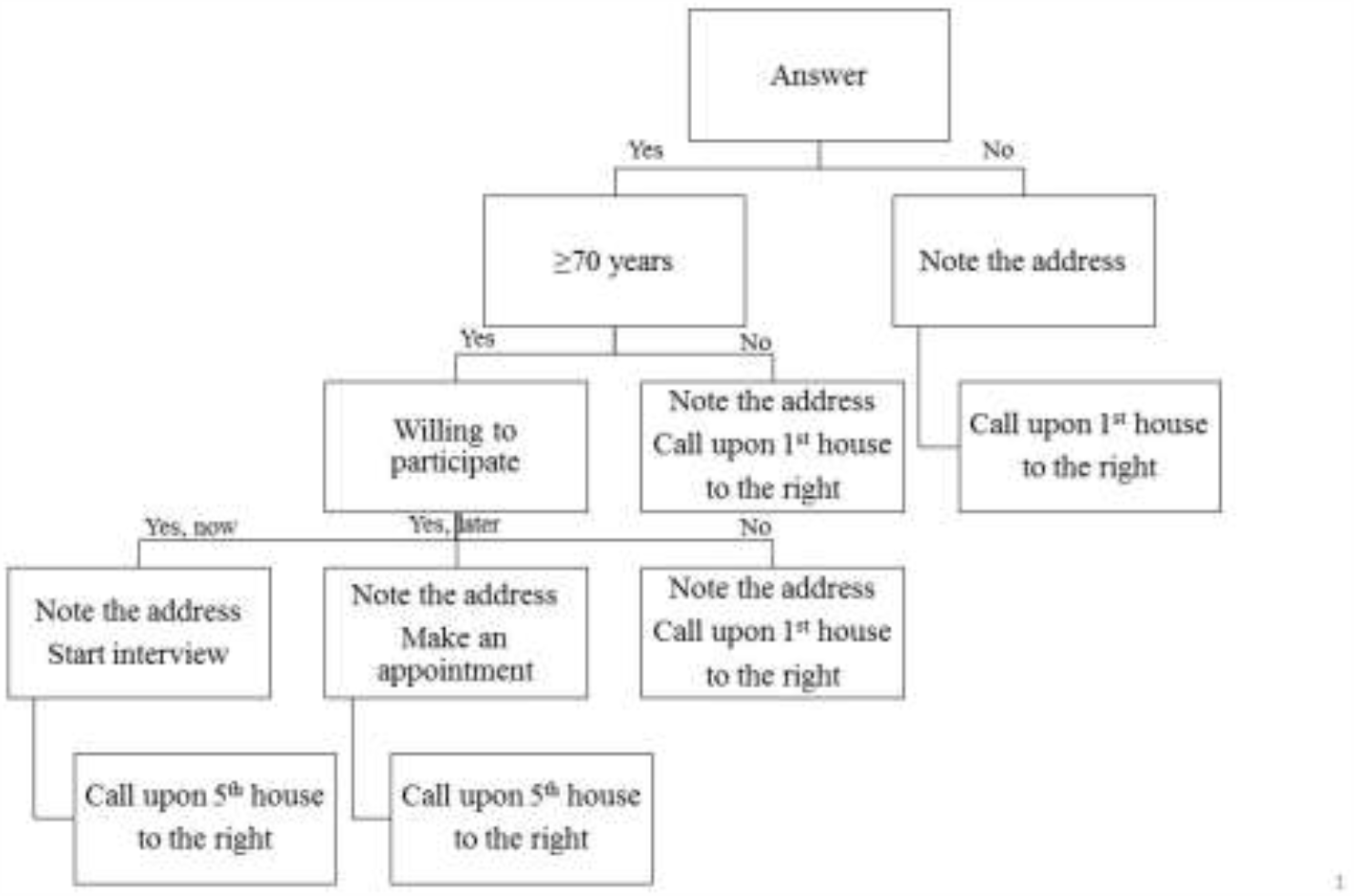
Flowchart random walk procedure^a^. ^a^Interviewers call upon every house on their route. After conducting an interview or making an appointment, they called on the 5^th^ house. At every crossing, they alternated between left and right. Every household was only invited once to participate. In nursing homes and assisted living facilities we randomly invited one person per unit using the Kisch selection method [32].

Face-to-face interviews were carried out in the participant’s home. Only one older adult per household could participate and proxy respondents were not allowed. If there was more than one eligible participant present, we used the late birthday selection rule to select our respondent [33]. Responses were collected using a tablet or computer. Interviews were conducted in Dutch, French or English. Participants received some chocolates or cookies to thank them for their participation.

### Ethical considerations

Ethical approval was obtained from the ethical committee of Ghent University/University Hospital (B670201837542). As is ethically sound in SV research [7], we presented the study as the “Belgian Survey on Health, Sexuality and Well-being”. All participants gave their informed consent before participating in the study. In line with the GDPR all data were pseudonymised. Documents containing personal data were kept separately from the answers to the questionnaire. Only the researchers working on the project had access to the personal data. All interviewers were trained in handling confidential information. In case of an acute dangerous situation (e.g., ongoing (sexual) violence), a multi-step safety procedure was in place in accordance to article 458 of Belgian criminal law. At the end of each interview, participants received a brochure containing contact details of several helplines.

## RESULTS

The data collection started on the 8^th^ of July 2019 and was stopped on the 12^th^ of March 2020, due to the COVID-19 pandemic and associated lockdown measures.

### Outcome random walk procedure

An overview of the random route outcome can be found in Table 1. We completed 513 interviews across Belgium, 116 (26%) were done by appointment, meaning that interviewers had to come back to the same household at least two times. Average duration of an interview was 54 minutes (range: 21-168 min, SD: 23 min). In order to complete 513 interviews, interviewers spent 320 days recruiting participants. They conducted 1.2 interviews a day on average. Yet, on 105 days no participants were found. In total 15,599 households were contacted, of which 1,805 (12%) were eligible to participate. Interviewers spent 515h 57min calling on doors. In order to complete one interview, interviewers called on average on 37 doors (SD: 31 doors) and spent 1h 28min (SD: 1h 37min). Participation rate in the different clusters ranged from 5% to 100%. Mean participation rate was 34% (SD: 19%).

**Table 1.**
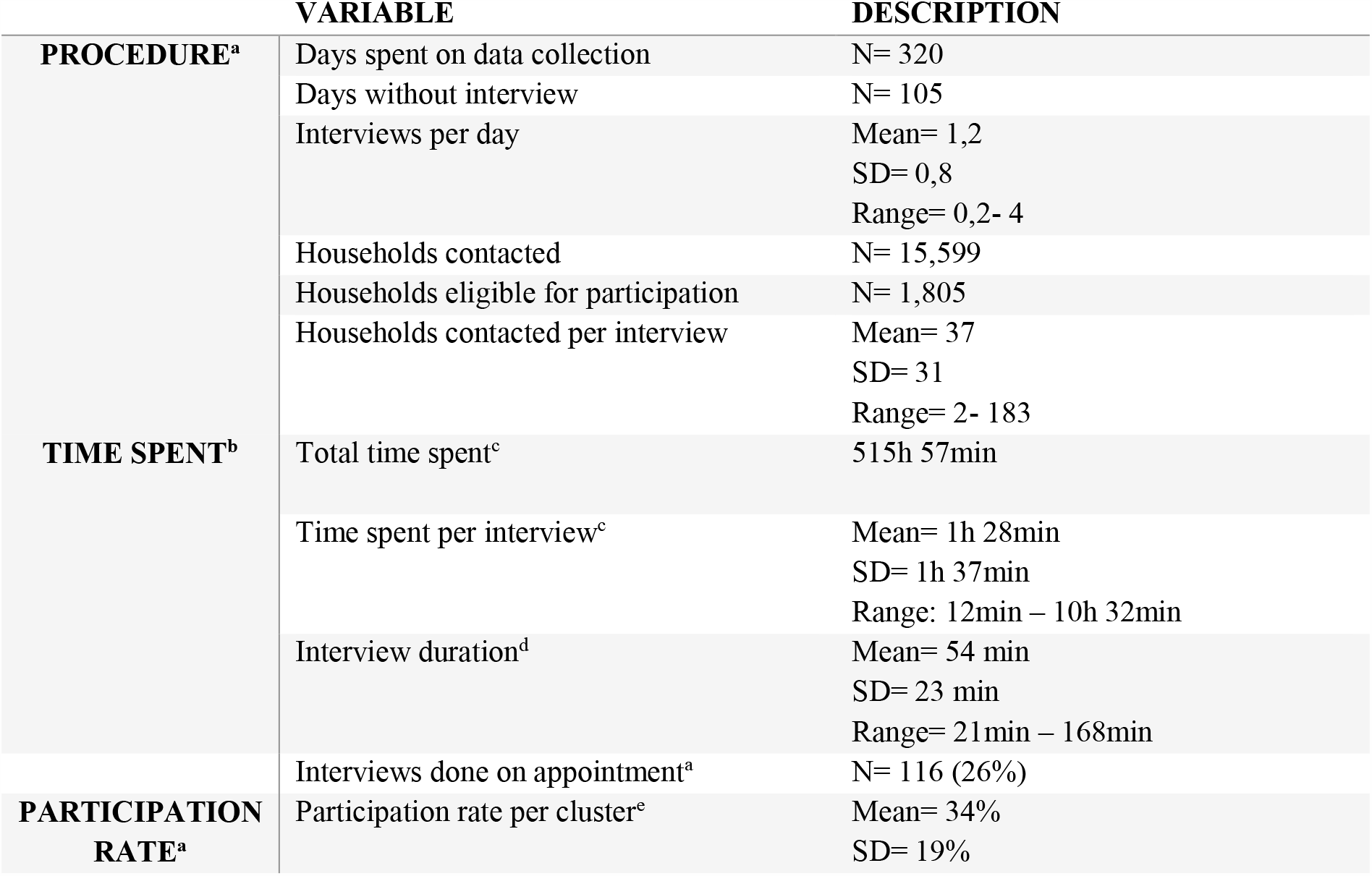
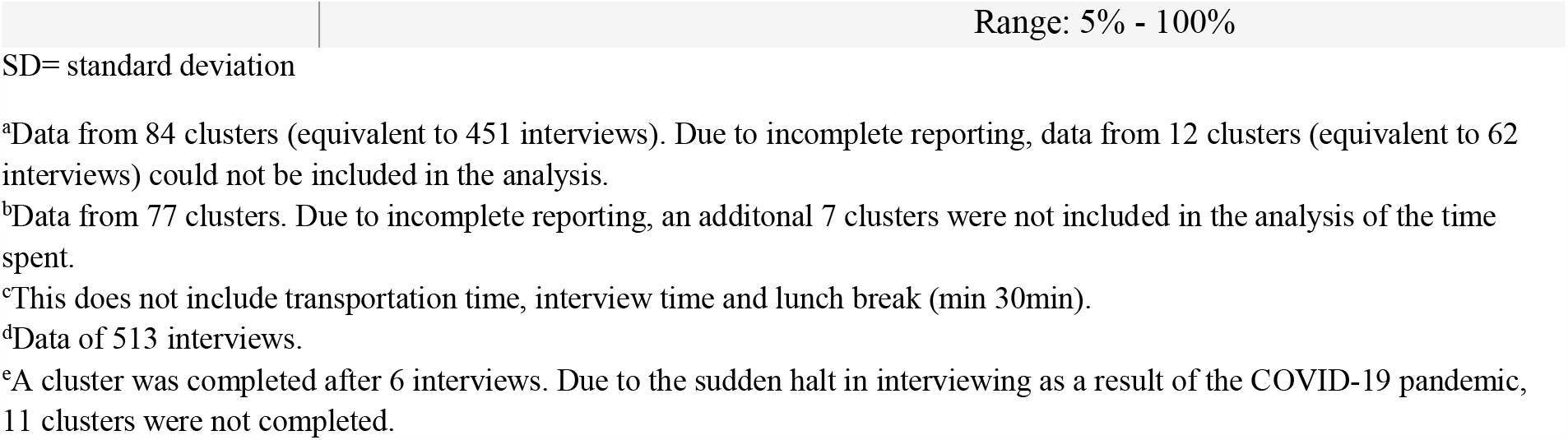
Outcome of the random walk procedure.

### Socio-demographics

Table 2 describes the sociodemographic characteristics of the study population in comparison to the Belgian population of 70 years and older. The random probability sampling method and random walk finding approach provided us with a study sample which is a valid representation of the Belgian older population. The distribution regarding gender, age and relationship status of our sample are comparable to those of the Belgian population of 70 years and older [1], however the number of nursing home residents is lower in our sample [34]. For education level we can only compare with the numbers of the Belgian population between 15 and 64 years old [1]. Regarding country of origin, our study reports on country of birth while the Belgian authorities describe nationality [1]. Considering sexual orientation, several participants experienced difficulties in understanding the different terms. Some heterosexual participants identified themselves as being “normal” and indicated “other” when answering this question, which could lead to an overestimation of people with non-heterosexual sexual orientation in our sample.

**Table 2:**
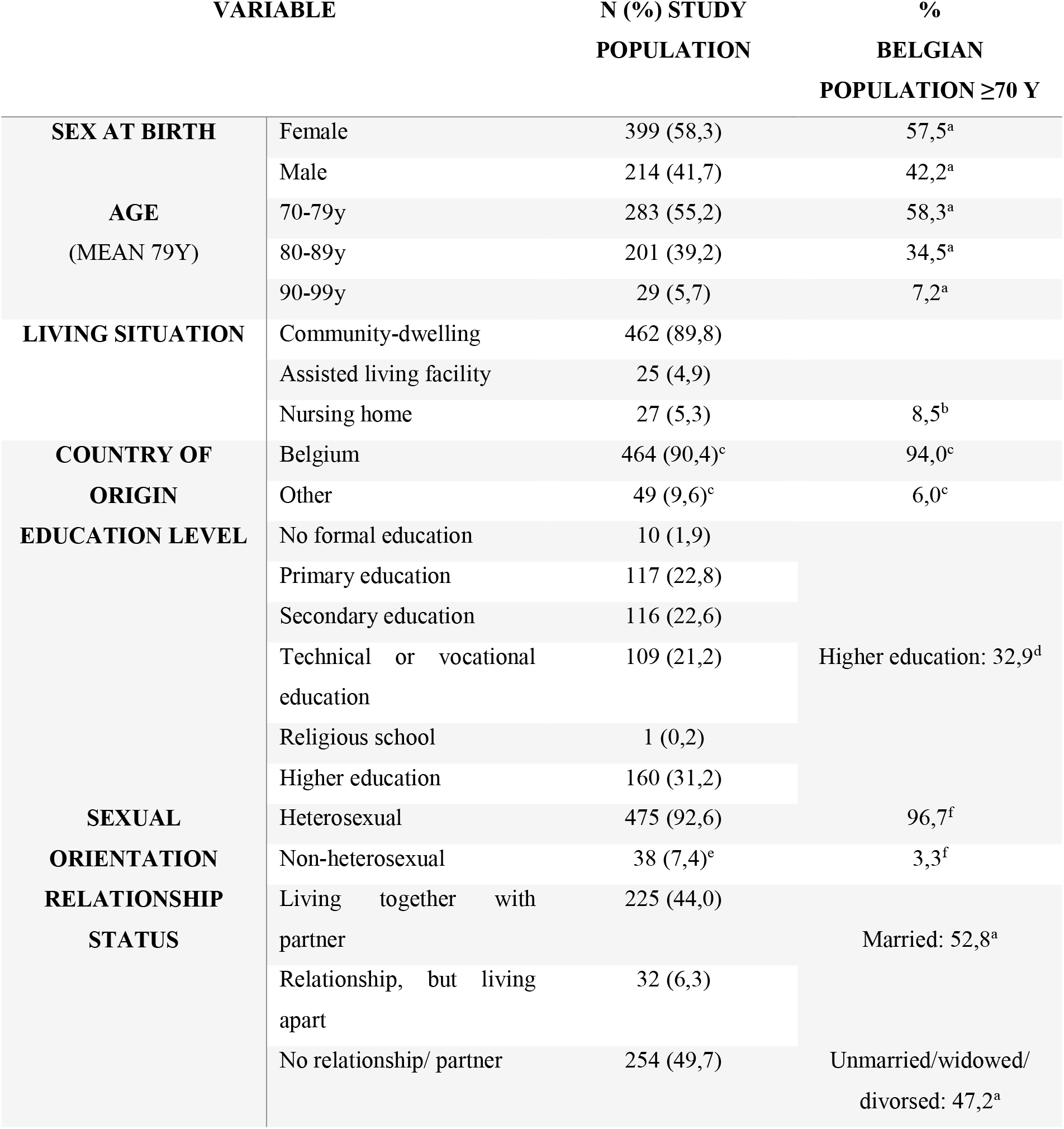
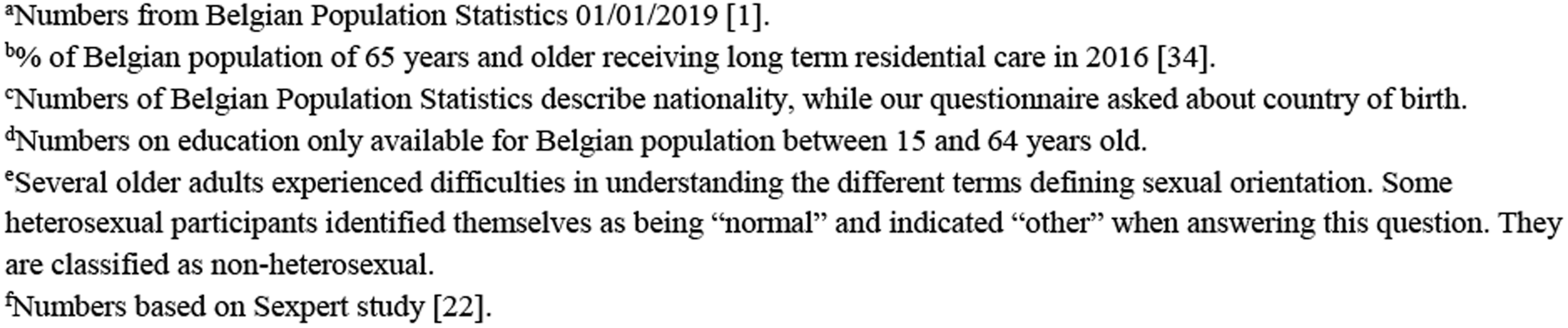
Sociodemographics of the study population (N=513)

| VARIABLE |  | N (%) STUDY<br>POPULATION | %<br>BELGIAN<br>POPULATION ≥70 Y |
| --- | --- | --- | --- |
| SEX AT BIRTH | Female | 399 (58,3) | 57,5<sup>a</sup> |
|  | Male | 214 (41,7) | 42,2<sup>a</sup> |
| AGE<br>(MEAN 79Y) | 70-79y | 283 (55,2) | 58,3<sup>a</sup> |
|  | 80-89y | 201 (39,2) | 34,5<sup>a</sup> |
|  | 90-99y | 29 (5,7) | 7,2<sup>a</sup> |
| LIVING SITUATION | Community-dwelling | 462 (89,8) |  |
|  | Assisted living facility | 25 (4,9) |  |
|  | Nursing home | 27 (5,3) | 8,5<sup>b</sup> |
| COUNTRY OF<br>ORIGIN | Belgium | 464 (90,4)<sup>c</sup> | 94,0<sup>c</sup> |
|  | Other | 49 (9,6)<sup>c</sup> | 6,0<sup>c</sup> |
| EDUCATION LEVEL | No formal education | 10 (1,9) |  |
|  | Primary education | 117 (22,8) |  |
|  | Secondary education | 116 (22,6) |  |
|  | Technical or vocational<br>education | 109 (21,2) | Higher education: 32,9<sup>d</sup> |
|  | Religious school | 1 (0,2) |  |
|  | Higher education | 160 (31,2) |  |
| SEXUAL<br>ORIENTATION | Heterosexual | 475 (92,6) | 96,7<sup>f</sup> |
|  | Non-heterosexual | 38 (7,4)<sup>e</sup> | 3,3<sup>f</sup> |
| RELATIONSHIP<br>STATUS | Living together with<br>partner | 225 (44,0) | Married: 52,8<sup>a</sup> |
|  | Relationship, but living<br>apart | 32 (6,3) |  |
|  | No relationship/ partner | 254 (49,7) | Unmarried/widowed/<br>divorced: 47,2<sup>a</sup> |

<sup>a</sup>Numbers from Belgian Population Statistics 01/01/2019 [1].
<sup>b</sup>% of Belgian population of 65 years and older receiving long term residential care in 2016 [34].
<sup>c</sup>Numbers of Belgian Population Statistics describe nationality, while our questionnaire asked about country of birth.
<sup>d</sup>Numbers on education only available for Belgian population between 15 and 64 years old.
<sup>e</sup>Several older adults experienced difficulties in understanding the different terms defining sexual orientation. Some heterosexual participants identified themselves as being “normal” and indicated “other” when answering this question. They are classified as non-heterosexual.
<sup>f</sup>Numbers based on Sexpert study [22].

### Interviewer’s feedback

During the debriefing meetings the interviewers rated the overall experience as rewarding. However, many interviewers experienced the door-to-door approach as challenging. Interviewers described feelings of disappointment, rejection and despair when the door-to-door approach was not successful. This led to a premature drop out of 21 interviewers (43%). As a consequence, a second pool of 8 interviewers was recruited and trained. The close follow-up was highly praised by the interviewers. The social media group in particular helped them to feel safe and stay motivated during the fieldwork. The group served as an accessible communication tool between the interviewers and the researchers and was used to exchange tips on interview techniques and practical questions. Besides the majority of interviewers having positive attitudes towards the social media groups, some interviewers felt pressured when reading success stories from others. Interviewers received positive feedback from respondents. Participants expressed their appreciations about the study and felt treated with respect. Participants openly discussed SH and SV with both same and opposite sex interviewers and 34% of victims disclosed their SV experience for the first time.

### Ethical and safety issues

During the fieldwork no acute dangerous situations were reported. Three interviewers became a victim of hands-off SV while interviewing. Every case was discussed in detail with the interviewers, the researchers and the coordinator of the study, upon which the interviewers found that no further actions needed to be taken. Two interviewers received an unwanted phone call by a respondent after the interview. As the informed consent form required both the name and signature of the interviewer, participants were able to retrieve the interviewers’ contact details via an internet search. They were not contacted again after having informed the callers to contact the coordinator instead of the interviewer.

## DISCUSSION

In this paper we outlined the methodology of a population-based study on SV in older adults in Belgium taking safety, ethical and privacy guidelines into consideration. We described how GDPR required us to change from a random sampling via the NR to a cluster random probability sampling and how we applied this procedure. Further, we explained how this change in sampling impacted the outcome of the study. In the discussion section we deliberate on the lessons learned and formulate recommendations for future studies.

When conducting this study, the research team had to overcome several challenges. First, we had to ensure older adults felt comfortable discussing SH and SV during a face-to-face interview. To maximise SV disclosure, the questionnaire built up towards the SV modules. Questions on SV and its consequences were phrased in a supportive and non-judgemental manner [7]. Since older adults may internalise the stereotypical societal image of ‘the asexual older adult’ [9-12] and talking about SH with older adults in considered inappropriate [13, 14], we took extra care to make the questionnaire acceptable to the target population. Our questionnaire was rigorously tested during a two-phase pilot study. This pilot study showed that allowing certain deviations off topic and discussing family life and SH at the beginning of the questionnaire, allowed participants to create a bond of trust with the interviewers. This helped participants to disclose SV later on. Furthermore, we carefully selected our interviewers and provided them with specialised training. Apart from introducing the questionnaire, the main goal of the training was to confront interviewers with their own attitudes, fears and stereotypes towards SV and older adults [7]. This helped them to minimize suggestion of judgement or blame when interviewing and to create a safe place to disclose SV. As a result, we received positive feedback from many participants. They felt they were treated respectfully and openly discussed SH and SV with both same and opposite sex interviewers, leading us to believe that same sex interviewers are not needed when interviewers are trained in non-judgemental communication. One third of SV victims disclosed their experiences for the first time.

Secondly, we had to ensure our data collection method did not endanger the safety of both participants and interviewers while being GDPR compliant. Since the opt-in procedure of the NR would lead to bias in participation and possible safety issues for respondents, we changed our sampling procedure to a cluster random probability sampling with a random walk finding approach. When going from door-to-door, protecting the safety of participants and interviewers was of the utmost importance. To that extent we took several actions: (1) every household could only be contacted once, (2) in nursing homes we contacted only one person per unit, (3) when conducting an interview or making an appointment, interviewers called on the 5^th^ house on their route, (4) only one person per household could participate,(5) proxy respondents were not allowed, (6) the study was introduced as the “Belgian Survey on Health, Sexuality and Well-being” and (7) in case of an acute dangerous situation, a multi-step safety procedure was in place in accordance with article 458 of Belgian criminal law. In addition, we provided interviewers with close support via email, telephone, social media and debriefing meetings. Based on our experiences, social media are adequate tools to facilitate the contact between the research team and the interviewers, but also between interviewers. Interviewers reported feeling safe and supported by the group while interviewing. However, some interviewers felt pressured by the progress of others. Therefore, we recommend always providing individual coaching for interviewers. During the fieldwork the safety of the participants was never endangered. In three out of 513 interviews, interviewers were exposed to hands-off SV. Two interviewers received an unwanted call of a participant. Close supervision of the interviewers ensured there were no major consequences. We recommend future studies on (sexual) violence to always take the WHO guidelines on violence research as a basis in designing the study. To protect the privacy of interviewers, we advise ethical committees to allow interviewers to sign the IC form with the name of the research group, without having to write down their own name.

The random probability sampling method and random walk finding approach provided us with a study sample which is a valid representation of the Belgian older population. The participation rate in our study was 34%, which is similar to the participation rates of previous studies on SH and elder abuse and neglect in Belgium and Europe [5, 22, 35], but lower than an Irish study on elder abuse and neglect using a multi-contacts approach [36]. Nonetheless, the random probability sampling method has proven to be time consuming and inefficient, especially given the specific safety constraints in SV research. As a result, we completed just 60% of the 845 planned interviews in approximately 68 working days. Only 12% of the 15,599 contacted households were eligible for participation. Time spent going from-door-to-door eclipsed time spent interviewing. In order to conduct one interview, interviewers had to contact on average 37 households and spent on average 1h 28min. As a result, the door-to-door approach was perceived as very challenging, leading to a premature drop-out of 21 interviewers and the need for additional recruitment and training. In addition, compared to random sampling via the NR, clustered sampling needs bigger samples to achieve the same level of representativeness. In our study, we needed to conduct twice as many interviews as originally planned, leading to a much higher costs. In order to keep conducting research on sensitive topics in older adults, it is vital that research institutions, donors, and policy makers convene to investigate how problems related to the GDPR can be solved in the future.

## CONCLUSION

Our study shows that doing research on SH and SV in older adults is feasible, taking safety, ethical and strict privacy guidelines into account. Older adults are willing to discuss SH and SV during a face-to-face interview by trained interviewers. The cluster random probability sampling method provides a good sampling frame to reach a diverse group of older adults resembling the actual older population. However, the method is inefficient and time-consuming resulting in a more expensive procedure. Adequate training and close supervision of interviewers is key to the success of the study. In order to keep conducting research on sensitive topics in hard to reach populations, it is vital that research institutions, donors, and policy makers convene to investigate how problems related to the GDPR can be solved.

## Data Availability

By request from the corresponding author.

## Acknowledgements

Prof. dr. Ines Keygnaert and Prof. dr. Christophe Vandeviver contributed equally to this work and are therefore to be regarded as joint last authors of this article. The authors want to thank Lotte De Schrijver and Joke Depraetere for their input during the questionnaire development. Also, we thank Prof. Dr. Olivier Degomme for his help in designing the sampling method of this study. Many thanks to our interviewers for their time and effort and to Dr. Howard Ryland for the language editing.

